# West Nile virus in Portugal

**DOI:** 10.1101/2021.02.02.21251021

**Authors:** José Lourenço, Sílvia C Barros, Líbia Zé-Zé, Daniel SC Damineli, Marta Giovanetti, Hugo C Osório, Fátima Amaro, Ana M Henriques, Fernanda Ramos, Tiago Luís, Margarida D Duarte, Teresa Fagulha, Maria J Alves, Uri Obolski

**Affiliations:** Department of Zoology, University of Oxford, United Kingdom; Instituto Nacional de Investigação Agrária e Veterinária, Virology Laboratory, Portugal; Centro de Estudos de Vectores e Doenças Infecciosas, Instituto Nacional de Saúde Doutor Ricardo Jorge, Portugal; Biosystems and Integrative Sciences Institute, Edificio TecLabs, Campus da FCUL, Portugal; Department of Pediatrics, Faculdade de Medicina da Universidade de São Paulo, Brazil; Laboratório de Flavivírus, Instituto Oswaldo Cruz Fiocruz, Brazil; Laboratório de Genética Celular e Molecular, Universidade Federal de Minas Gerais, Brazil; Instituto de Saúde Ambiental, Faculdade de Medicina da Universidade de Lisboa, Portugal; School of Public Health, Faculty of Medicine, Tel Aviv University, Israel; Porter School of the Environment and Earth Sciences, Faculty of Exact Sciences, Tel Aviv University, Israel

**Keywords:** west nile virus, epidemiology, climate, modelling, portugal

## Abstract

West Nile virus (WNV) causes outbreaks with sustained spillover to humans in many European countries. Despite Portugal’s Mediterranean climate being adequate for WNV transmission, only four human infections have been detected there so far. Here, we offer an historical account of past WNV circulation and develop new, climate-driven insights on the geo-temporal suitability for WNV transmission in Portugal. WNV and vector related literature and database reviews were performed in the context of Portugal covering the time period 1966-2020, and local climate data were used to estimate WNV transmission suitability for the period 1981-2019. Reviewed data demonstrate that WNV-compatible vectors are abundant across the entire country, while molecular and serological evidence for WNV circulation has mostly been associated with the southern districts. Our estimated WNV transmission suitability was found to support geographical differences in transmission potential that favour the southern districts, with an increasing trend over the past forty years due to climate change. Empirical and theoretical evidence supports WNV circulation in Portugal, but it remains unclear whether the virus is endemic or sporadically transmitted. Given the recent public health emergencies related to WNV in other European countries and the findings herein described in relation to Portugal, our study supports a timely change towards a local WNV active surveillance.

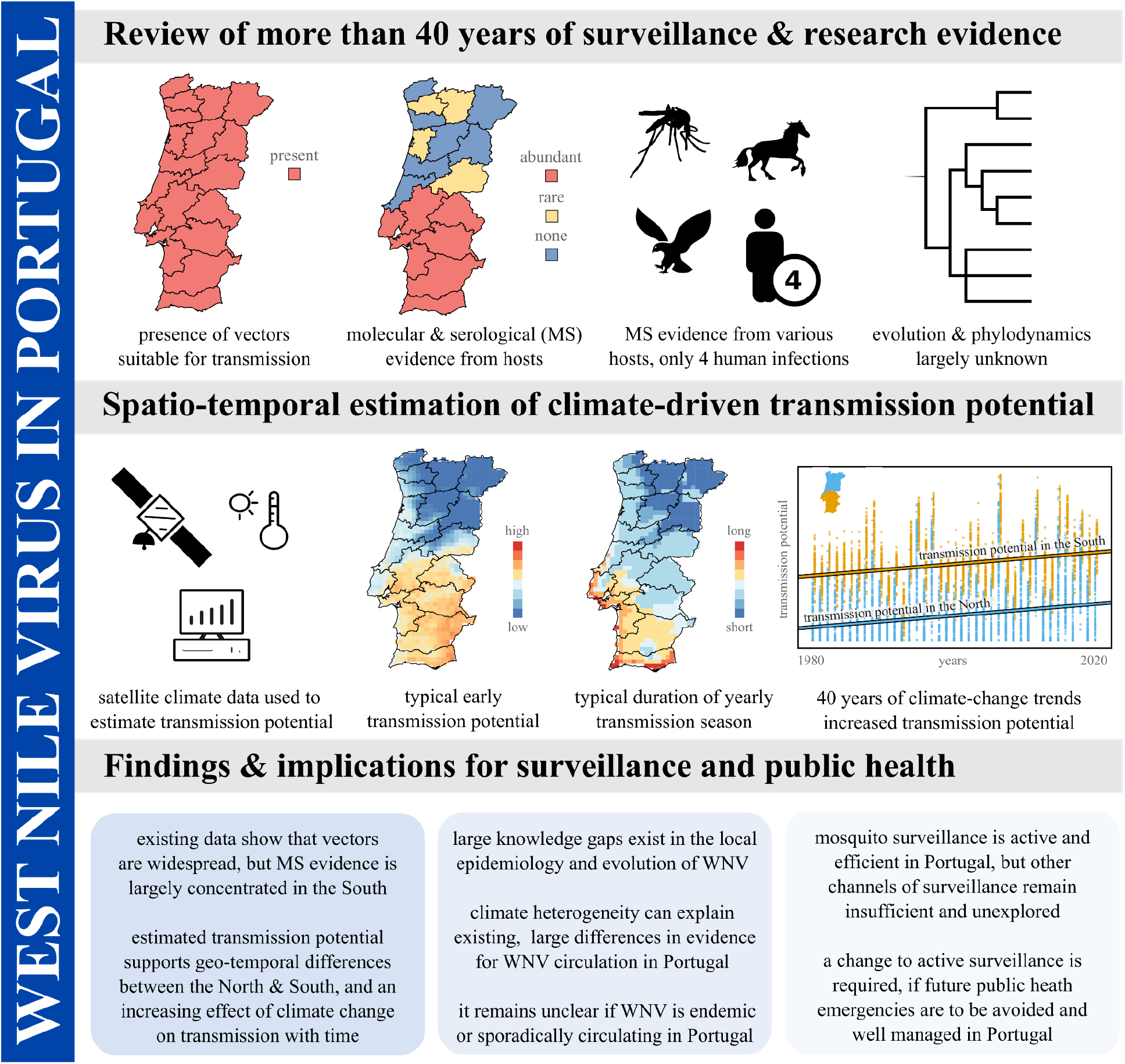

## 1 Introduction

West Nile virus (WNV) is an RNA mosquito-borne virus of the Flaviviridae family, first identified in 1937 in the West Nile district of Uganda. WNV ecology is characterised by a zoonotic transmission cycle maintained between a large number of mosquito and wild avian species. Occasional viral spillover can occur from this cycle to humans as well as domesticated and wild mammals [1–3]. From such incidental hosts, humans and equines feature predominantly in epidemiological data given a clear West Nile fever clinical profile, with equines often serving as a sentinel species in many countries. Contrary to most avian species, mammals are inefficient amplifier hosts, and hence can not establish mosquito-mammal transmission cycles [3–5]. Approximately 80% of infections in humans are asymptomatic while the rest may develop mild or severe disease of neuroinvasive nature and potentially death [6]. There are currently no licensed vaccines nor antiviral treatments for humans [3,7], but four licensed vaccines are on the market for use in equines [7].

Global trends in climate change and human factors continue to favour the dispersal of mosquito species into new regions, and the long-distance movement of infectious hosts across the globe. Both factors promote the introduction and epidemic activity of mosquito-borne viruses such as WNV into previously unaffected areas [8–11]. Some of the most relevant mosquito-borne examples of the past 10 years include the introduction of chikungunya and Zika viruses into the Americas [12,13], as well as the resurgence of yellow fever virus in Brazil and regions of Africa [14,15].

In the 20th century, WNV was mostly reported to circulate in Israel and some African countries [16]. More recently, in 1999, it gained international notoriety after its introduction into the United States of America (USA, New York), from where it quickly became endemic in most of the USA and Canada [17]. Concurrently and in the following two decades, epidemic activity increased in regions of Europe, the Middle East and Russia [11,16]. Presently, WNV epidemiology shows large spatio-temporal heterogeneities across countries and continents, but much of that variation is not well understood. However, given its well documented effects on the life-cycles and dispersal of avian and mosquito species, climate has been acknowledged as a major factor influencing local WNV epidemic activity, dispersal and persistence [3,18,19].

The recent epidemiological history of WNV in Europe is characterised by increasing epidemic activity in countries including France, Italy, Greece, Hungary, Romania, Bulgaria, Serbia and Ukraine [20,21]. In 2018, the continent reported its largest epidemic ever across many countries, exceeding cumulative infections since 2010 [22]. In 2020, the first autochthonous human infections were reported in the Netherlands [23], and Spain had its largest epidemic to date including regions previously not affected [24,25]. Compared to Spain and other countries in the Mediterranean basin, Portugal remains an outlier in so far having reported only four, serologically confirmed human infections in the past 40 years.

Routine laboratory diagnostics in Portugal were established in 1996, and are currently managed by the Centro de Estudos de Vectores e Doenças Infecciosas (CEVDI, Center for Vectors and Infectious Diseases Research) of the Instituto Nacional de Saúde Dr. Ricardo Jorge (INSA, National Health Institute Dr. Ricardo Jorge) and by the Instituto Nacional de Investigação Agrária e Veterinária (National Agrarian and Veterinary Research Institute, INIAV) in human and animals, respectively. Notification of WNV-related disease is currently mandatory in Portugal. Being an European Union (EU) member state, Portugal reports outbreaks of equine WNV neurological disorders to the European Animal Disease Notification System [26], and annual reports for the EU region are published by the European Food Safety Authority [27] and European Centre for Disease Prevention and Control (ECDC) [28].

Since 2008 the country has been performing extensive mosquito surveillance under REVIVE (REde de VIgilância de VEtores, surveillance network for vectors [29,30]). A multitude of national agencies within the Ministry of Health are involved, including the General Directorate of Health (DGS), INSA, the five Regional Health Administrations (Algarve, Alentejo, Lisboa e Vale do Tejo, Centro and Norte), and the non-continental agencies the Institute of Health Administration of Madeira and the Regional Health Directorate of the Azores. REVIVE is responsible for the nationwide surveillance of the most significant hematophagous arthropods for national public health (mosquitoes, ticks, and sandflies) including the cataloguing of extant species and their population sizes. Surveillance of invasive mosquito species, such as *Aedes aegypti* and *Aedes albopictus*, and screening (e.g. PCR) of a multitude of field-collected mosquito-species for viruses is also regularly performed. According to International Health Regulations, airports, ports, storage areas, and specific border regions with Spain are regularly monitored throughout the year according to the needs of local and regional authorities. Reports from REVIVE are provided regularly to national agencies across the country [29], and to the European Network for Arthropod Vector Surveillance for Human Public Health, a network for data sharing on the geographic distribution of arthropod vectors, transmitting human and animal disease agents (originally known as VBORNET, and VectorNet since 2014 [31]).

WNV surveillance in Portugal remains passive and mostly reactive to occasional reports of clinically compatible events in equines and humans. However, many local clinicians and veterinarians lack awareness of the importance of WNV screening in compatible disease cases. As such, and since infections tend to be asymptomatic or mild, it is possible that only a very small percentage of all infections end up being screened and therefore accounted for. Detection of WNV RNA in serum, cerebrospinal fluid or urine offer significant technical challenges in both humans and equines when assessed substantially after the start of the symptomatic period. In Portugal, WNV diagnosis in humans is typically requested following the failure of all other disease compatible diagnoses, commonly days or weeks after symptoms onset and consequently after the viraemic phase. The reactive nature of the surveillance system also implies that increased awareness and screening requests for humans and equines tend to rise around areas with recent positive cases, but only for very short time windows which remain insufficient for long-term risk and epidemiological assessment. These aforementioned realities allow for systematic gaps in current knowledge regarding the local epidemiology of WNV, hindering important conclusions such as whether or where the virus is endemic.

In this article, we review and describe the international and Portuguese literatures and official data sources (1966-2020), further providing a theoretical geo-temporal assessment of WNV transmission potential in Portugal (1981-2019). We find a clear north-south dichotomy in existing evidence for the circulation of WNV in Portugal, although high resolution data on WNV-capable mosquito species suggests transmission would be possible across the entire country. Our theoretical assessments on transmission potential suggest that the south of the country presents climate-driven seasonal signatures that justify WNV circulation to be restricted to that region. Using climate data of the past four decades, we also quantify long-term trends in climate and their possible historical effects on the local WNV transmission potential. In light of our conclusions and the recent increase in epidemic behaviour of WNV in the European region, we advocate for a more active surveillance of the virus in Portugal, in which ongoing circulation may soon develop into a public health emergency.

## 2 Material and Methods

### 2.1 Literature reviews

We queried PUBMED for articles related to WNV and Portugal using the query “((west nile virus) AND (Portugal)) OR (vírus nilo AND Portugal)” in all fields; thus leaving the query non-specific enough to capture a wide range of studies, and searching for articles both in Portuguese and English. The query returned 43 hits as of March 2021, from which 17 did not relate directly to the circulation of WNV or its vectors in the country. A second literature search was performed manually among Portuguese academic journals and PUBMED. References within the studies found in our two searches were also scanned for potential studies related to WNV circulation in Portugal. We also scanned the GenBank nucleotide database for existing WNV sequences from Portugal, searched with the query “((west nile virus) AND (Portugal)) OR ((WNV) AND (Portugal))” OR (vírus nilo AND Portugal). The query returned 70 hits as of March 2021, of which 62 did not relate to WNV genomic regions.

We provide the results of our literature review in the form of tables in the main text. Studies found in the PUBMED and manual searches are grouped by content type in **Table 1**. All evidence related to the presence of WNV in Portugal across all studies found is summarized in **Table 2** (also available in a permanent figshare repository [32]). Studies presenting time series of relevant mosquito species in Portugal are summarized in **Table 3**. All publicly available WNV genetic sequences obtained in Portugal are listed in **Table 4**.

**Table 1.** Studies including information on WNV or its vectors in Portugal.

| Group | PUBMED [in English] | Manual [in English & Portuguese] |
| --- | --- | --- |
| WNV in animals, in Portugal<br>(serology / viral isolation / VNT) | [52,53,58,61,72] | [52,54,57,73] |
| WNV in vectors, in Portugal<br>(viral isolation) | [55,59,60,72,74–76] | [57] |
| WNV in humans, in Portugal<br>(serology / viral isolation / VNT) | [65,72] | [56,64] |
| WNV vectors in Portugal but no<br>WNV research | [30,67,77–83] | [84] |
| WNV modelling including regions<br>of Portugal | [85–87] |  |
| WNV phylogenetics in Portugal | [76] |  |

**Table 2.** Sources including past evidence for WNV circulation in Portugal.

| Sample | Type | N | Region | Period | Study |
| --- | --- | --- | --- | --- | --- |
| Equine | VNT | 7/24 | Aljustrel | Summer 1970 | [54] |
| Human | HI | 38/1649 | Country wide | 1969 - 1973 | [56]♦ |
| Mammal | HI | 16% | Portalegre, Évora, Beja | Sep 1966 - Feb 1967 | [53]♦ |
| Mammal | HI | 3% | Portalegre, Évora, Beja | Sep 1966 - Feb 1967 | [53]♦ |
| Vector | PCR | pool | Ancão, Almancil | Jul 2004 | [59]♦ |
| Vector | Isolation | pool | Barragem do Roxo | Sept 1969 | [55] |
| Human | Serology | 2 | Faro | Jun - Jul 2004 | [63] |
| Human | Serology | 1 | Setúbal | Jul 2010 | [64] |
| Equine | VNT | 40/1313 | Santarém, Aveiro, Portalegre, Évora, Setúbal, Algarve | 2004 - 2010 | [58]■ |
| Avian | VNT | 23/116 | Vila Real, Lisboa, Setúbal, Algarve | 2004 - 2010 | [58]■ |
| Equine | VNT | 22/23 | Loulé | Aug - Nov 2015 | [61]■ |
| Equine | VNT | 1/1 | Faro | Aug - Oct 2015 | [61]■ |
| Equine | VNT | 1/1 | Olhão | Sept 2015 | [61]■ |
| Equine | VNT | 8/9 | Lagos | Sept - Oct 2015 | [61]■ |
| Equine | VNT | 4/4 | Alcácer do Sal | Sept - Oct 2015 | [61]■ |
| Equine | VNT | 1/1 | Arronches | Oct 2015 | [61]■ |
| Equine | VNT | 1/1 | Alpiarça | Oct 2015 | [61]■ |
| Equine | VNT | 2/2 | Loulé | Feb - Oct 2016 | [61]■ |
| Equine | VNT | 1/1 | Lagos | Jul - Dec 2016 | [61]■ |
| Equine | VNT | 3/3 | Silves | Oct - Dec 2016 | [61]■ |
| Equine | VNT | 1/1 | Odemira | May 2016 | [61]■ |
| Equine | VNT | 1/1 | Salvaterra de Magos | Jun - Dec 2016 | [61]■ |
| Equine | VNT | 2/2 | Portalegre | Sep - Nov 2016 | [61]■ |
| Equine | VNT | 1/1 | Alter-do-chão | Oct 2016 | [61]■ |
| Equine | VNT | 1/1 | Benavente | Nov 2016 | [61]■ |
| Equine | VNT | 1/1 | Elvas | Nov - Dec 2016 | [61]■ |
| Equine | VNT | 1/1 | Beja | Nov - Dec 2016 | [61]■ |
| Vector | PCR |  | Ria Formosa | Jul 2004 | [60] |
| Avian | HI | 1/4 | Parque Nacional da Peneda-Gerês | 1999 - 2002 | [57]◆ |
| Avian | HI | 1/19 | Parque Natural do Vale do Guadiana | 1999 - 2002 | [57]◆ |
| Avian | HI | 3/44 | Parque Ecológico de Monsanto | 1999 - 2002 | [57]◆ |
| Avian | HI | 2/12 | Parque Ecológico de Monsanto | 1999 - 2002 | [57]◆ |
| Avian | HI | 1/4 | Parque Nacional da Peneda-Gerês | 1999 - 2002 | [57]♦ |
| Avian | HI | 1/4 | Parque Ecológico de Monsanto | 1999 - 2002 | [57]♦ |
| Avian | HI | 2/6 | Parque Nacional da Peneda-Gerês | 1999 - 2002 | [57]♦ |
| Avian | HI | 1/3 | Reserva Natural de Santo André | 1999 - 2002 | [57]♦ |
| Avian | HI | 1/3 | Reserva Natural de Santo André | 1999 - 2002 | [57]♦ |
| Avian | HI | 3/13 | Parque Nacional da Peneda-Gerês | 1999 - 2002 | [57]♦ |
| Equine | Serology | 3/41 | Santarém | 1999 - 2002 | [57]♦ |
| Human | VNT | 1 | Almancil | Jul 2015 | [65] |
| Equine | Serology | 1/18 | Loulé | Aug 2015 | [62] |
| Equine | Serology | 1/54 | Faro | Aug 2015 | [62] |
| Equine | Serology | 2/10 | Loulé | Aug 2015 | [62] |
| Avian | ELISA | 8/123 | Reserva Natural de Santo André, Tagus estuary, Sado estuary | 2006, 2008 - 2010 | [73]♦ |
| Avian | ELISA | 5/46 | Reserva Natural de Santo André, Tagus estuary, Sado estuary | 2006, 2008 - 2010 | [73]♦ |
| Avian | ELISA | 9/213 | Reserva Natural de Santo André, Tagus estuary, Sado estuary | 2006, 2008 - 2010 | [73]♦ |
| Equine | ELISA / PCR | 23/165 | Loulé, Portalegre, Silves, Alter-do-Chão, Lagoa, Elvas, Lagos, Benavente, Alcácer do Sal, Alcáçovas, Samora Correia, Albufeira, Escalos de Cima-Castelo Branco, Comporta | Aug 2016 - Sep 2020 | This study |

■: Also IgM and prE-IgG serology, VNT only used when confirmed serology; ♦: positive, implying Flavivirus cross-reactivity, but no WNV confirmed by other means; HI: hemagglutinin inhibition; VNT: virus neutralization test; PCR: polymerase chain reaction; ELISA: enzyme-linked immunosorbent assay.

**Table 3.** Articles including time series of mosquito population dynamics in Portugal.

| Source | Region, period | Study |
| --- | --- | --- |
| *Aedes berlandi*, *Aedes caspius*, *Anopheles atroparvus*, *Anopheles claviger*, *Culex hortensis*, *Culex laticinctus*, *Culex mimeticus*, *Culex* | Alqueva, monthly, 2004-2005, 2007 | [74] |
| *pipiens*, *Culex theileri*, *Culex univittatus*, *Culiseta annulata*, *Culiseta longiareolata*, *Culiseta sunbochrea* |  |  |
| *Aedes caspius*, *Aedes detritus*, *Anopheles atroparvus*, *Culex pipiens*, *Culex theileri*, *Culex univittatus*, *Culiseta annulata*, *Culiseta longiareolata*, *Culiseta subochrea* | Comporta, monthly, 2004-2005 | [74] |
| *Aedes caspius*, *Aedes detritus*, *Anopheles atroparvus*, *Anopheles algeriensis*, *Coquillettidia richiardii*, *Culex modestus*, *Culex pipiens*, *Culex theileri*, *Culex univittatus*, *Culiseta longiareolata* | Algarve, monthly, Jun Sep 2007 | [84] |
| *Culex pipiens*, *Ochlerotatus caspius* | Sado estuary, Ria Formosa, monthly, May Oct 2005, 2006 | [84] |
| *Culex pipiens* | Entire country, 2016 - 2019 | REVIVE [29] |

**Table 4.** Existing WNV genetic sequences from Portugal.

| Location | Host | Genomic region | Year | Accession ID |
| --- | --- | --- | --- | --- |
| Algarve | Mosquitoes | NS5 | 2004 | AJ965631 |
| Algarve | Mosquitoes | NS5 | 2004 | AJ965627 |
| Algarve | Mosquitoes | Polyprotein precursor | 2004 | AJ965630 |
| Algarve | Mosquitoes | Polyprotein precursor | 2004 | AJ965629 |
| Algarve | Mosquitoes | Polyprotein precursor | 2004 | AJ965628 |
| Algarve | Mosquitoes | Polyprotein precursor | 2004 | AJ965626 |
| Alentejo | Mosquitoes | Polyprotein precursor | 1969 | AM404308 |
| Alentejo | Mosquitoes | Env | 1969 | AY727828 |

### 2.2 Mosquito data

We used mosquito data collected by the REVIVE initiative in Portugal [29]. Data included mosquito counts (adult and immature stages) for *Culex pipiens* per county in Portugal during the period 2017-2019. The REVIVE data is in the public domain [29].

### 2.3 Transmission suitability

The basic reproduction number R_0_ of WNV in the zoonotic reservoir can be summarised as the product of two factors: the number of adult female mosquitoes per host *M*, and the transmission potential of each adult female mosquito P(i.e. *R*_0_ *= MP)*. Both *M* and P oscillate in time, driven by the influence of multiple eco-climatological factors. *M*, the ratio between the population size of mosquitoes *V* and hosts *N (M = V/N)*, depends on the local effects of climate on mosquito survival and reproduction, affecting *V* [33], as well as on the reproduction and migratory behaviours of the avian species, affecting *N* [34]. For most epidemiological contexts and pathogens, high resolution data on either *V* or *N* is often lacking. The transmission potential *P* depends on viral, mosquito and host factors (e.g. incubation periods, lifespans, etc), some of which depend on local climate variations [33,35,36]. Since adequate geo-temporal data for *V* and *N* is generally difficult to obtain, many recent theoretical epidemiology studies resort to estimating the transmission potential of mosquito-borne viruses using ‘suitability indices’. These indices are functions of the parameters that compose either *M* or *P* or both, and generally depend on covariates such as climatic variables, vegetation indices, host demography, mosquito and disease incidence reports, etc [36–38]. While a suitability index is an incomplete measure of the real transmission potential of a mosquito-borne virus, much work has been done to demonstrate the potential of an index to capture the geo-temporal dynamics of the number of reported mosquito-borne infections in host species [36,39,40].

In this study, we assessed local WNV transmission potential by estimating one of such suitability indices - termed the index *P* (the transmission potential of each adult female mosquito from *R*_0_ = *MP*) The theory and practice of *P* has been previously described in full by Obolski et al., that also introduced the MVSE R-package capable of estimating *P* given some epidemiological priors and climatic data [36]. The index *P* has been used in several studies focusing on the population dynamics of the dengue, chikungunya, and Zika viruses (e.g. [36,39,41,42]). Recently, the index was also successfully used to estimate the transmission potential of WNV in Israel dependent on local relative humidity and temperature variables [43].

In that study, *P* was interpreted as a proxy for the risk of spillover to the human population given that it measured the WNV transmission potential by single adult female mosquitoes in the animal reservoir. In the current study, we use the same approach as applied to Israel, informed by the epidemiological priors applied in that study (see Table in main text of [43]), which relate to spp. *Culex*, WNV and a theoretical, average bird species.

To inform the estimation of the index *P*, climatic data was downloaded from the Copernicus platform at www.copernicus.eu. We used the available dataset “essential climate variables for assessment of climate variability” [44], which includes climatic variables at a time resolution of 1 month (1981-2019), and gridded spatial resolution of 0.25°x0.25°. In the current study we used MVSE R-package version 1.01r [45], which uses temperature, relative humidity and precipitation as input data. For each latitude-longitude Portuguese data point in the Copernicus dataset, we estimated the index *P* as described by Obolski et al. for Brazil, Honduras and South America [36]. The resulting geotemporal estimations, as presented in the main text, are made available as a comma separated file in a permanent figshare repository [32].

### 2.4 Analysis of peak suitability timing and size

Peaks in the monthly index *P* time series 1981-2019 per county were found by the maximal value and month where it occurred, using a circular mean to calculate the average time point of the year for the north and south regions, independently. The distribution of values throughout the years were then compared between regions with a circular ANOVA yielding a *P* = 0. 005. Circular data was handled with the R-package ‘circular’ [46]. Mean peak values were calculated across counties for each year, using Welch’s t-test to compare north and south regions yielding a *P* = 1 ⨯ 10^−16^.

### 2.5 Analysis of long-term climatic and suitability trends

We applied linear quantile mixed models to the estimated index *P*, temperature, precipitation and humidity between 1981 and 2019, for which the Copernicus climate dataset was available. Our data contained yearly median values of the mentioned variables for each Portuguese county. Since the data included highly irregular points, which might skew standard linear regression models, we chose to use the more robust quantile regression using the median (τ = 0. 5). Furthermore, as different regions had repeated measures, we employed a mixed-effects quantile regression (using the ‘lqmm’ R-package [47]). Our models contained a random intercept term for each region, a fixed effect dummy variable coding for north versus south, and a continuous variable representing the time unit (year). We tested the interaction between the year and north-south terms and found it statistically significant (*p* < 0. 01) in all models except when index *P* was the dependent variable (*p* >0.7) for which the term was dropped. All the other fixed effects were statistically significant for all models (*p* < 0. 001) and are summarized in **Table 5**.

**Table 5.** Statistical output from the linear quantile mixed modelling.

| Dependent variable | Year slope (95% CI) | South intercept (95%CI) | South*year slope (95% CI): |
| --- | --- | --- | --- |
| Index P | $9.8 \times 10^{-4}$<br>($9.4 \times 10^{-4}$, $10^{-3}$) | 0.082<br>(0.077, 0.088) | ----- |
| Humidity (relative, %) | - 0.07<br>(- 0.08, - 0.06) | - 65<br>(- 94, - 36) | 0.03<br>(0.015, 0.045) |
| Temperature<br>(Celsius) | 0.04<br>(0.04, 0.04) | 9.6<br>(5.3, 13.9) | - 0.003<br>(- 0.005, - 0.001) |
| Precipitation<br>(m day<sup>-1</sup>) | - 1.2 × 10<sup>-6</sup><br>(- 1.8 × 10<sup>-6</sup>, - 6.6 × 10<sup>-7</sup>) | - 0.013<br>(- 0.015, - 0.014) | 6 × 10<sup>-6</sup><br>(5 × 10<sup>-6</sup>, 7 × 10<sup>-6</sup>) |

### 2.6 Analysis of long-term suitability periodicity

Detrending and wavelet analysis followed the workflow of Damineli et al. [48]. Detrending with a Maximal Overlap Discrete Wavelet Transform was performed in a multiresolution analysis with the R-package ‘wavelets’ [49]. Significant periods of the continuous wavelets were assessed considering the spectrum of an autoregressive model of order 1, as a null model using the R-package ‘biwavelet’ [50].

### 2.7 Equine WNV surveillance data 2016-2020

Between 2016 and 2020 the virology laboratory of INIAV performed molecular and serology testing in 165 horses from several regions of the country, reacting to local suspected neuroinvasive disease. RNA was extracted by using the BioSprint 96 workstation with the MagAttract 96 cador pathogen kit, according to manufacturer’s instructions (Qiagen, Hilden, Germany). The samples were screened for WNV by real-time reverse transcriptase PCR (RT-qPCR) targeting the NS2A gene [51]. Equine sera were tested for WNV-IgM antibodies by capture ELISA (ID Screen West Nile IgM Capture ELISA, IDVET, Montepellier, France). Twenty three horses were diagnosed as WNV-positive on the basis of IgM positivity, with one horse being also positive by RT-qPCR (July 2020, Escalos de Cima-Castelo Branco, center Portugal). Four of eight horses that were followed for clinical progression died or were euthanized. These results are summarized in **Table 2**, including the time and region of sampling.

## 3 Results

### 3.1 Evidence for West Nile virus circulation in Portugal

The earliest evidence of WNV circulation in Portugal came from serological surveys in the south of the country in 1966-1967, reporting high titres of hemagglutination-inhibiting antibodies and neutralizing capacity against WNV in animal serum [52,53]. Such findings led to several initiatives in 1969 to create institutional links between the national health and veterinary medicine services with the purpose of seeding future studies and raising awareness to the virus [54]. Some of the earlier initiatives included the capture and testing of animals (e.g. equines, rodents, birds, mosquitoes) and serological testing of human serum [53–56], eventually leading to the first viral isolation from the mosquito species *Anopheles maculipennis* [55].

After a period of three decades with seemingly no research output in the country, further evidence for the circulation of WNV in Portugal accrued from several studies. Between 1999 and 2002, Formosinho and colleagues reported serological data from several avian species sampled across natural reserves and from equines sampled in the district of Santarém [57]. A few studies in the period 2004-2010 also reported positive findings in several avian species [58], mosquitoes [59,60], and equines [58]. More recently, between 2015 and 2020, following reports of several clinically compatible equine outbreaks in the south of the country, a number of equines were found serologically positive to the virus in the Alentejo and Algarve [61,62]. Some of this evidence is reported for the first time in this study (see **2 Material and Methods** for details, and **Table 2** for summary).

Although such evidence supports a long history of local circulation in animals and mosquitoes, autochthonous WNV infections in humans in Portugal were only first reported in 2004. This was the case of two Irish bird-watchers found positive for both WNV IgG and IgM antibodies. They visited Ria Formosa, a marshland rich in endemic and migratory birds in the Algarve between the 26th June and 10th of July [63]. Two studies published thereafter were able to detect WNV by RT-PCR in pools of mosquitoes collected near the putative region of infection that year [59,60]. A third case was reported in July 2010 in the region of Setúbal, from a local rural resident with no history of international travel [64]. The resident tested positive for both WNV IgG and IgM antibodies in the weeks following the clinical diagnosis. Reactive mosquito collection performed around the patient’s residence between the 13-18th of July revealed species suited for WNV transmission (e.g. *Culex pipiens*), although all mosquito pools tested negative by RT-PCR [64]. Two months later, two equine cases were notified about 4 kilometers away from the reported human case. The most recent human infection was reported in a rural area of Almancil (Algarve) in 2015 in a patient testing positive for both WNV IgG and IgM antibodies and viral neutralization, with no record of international travel or history of vaccination against flaviviruses [65]. In August and September of the same year several outbreaks in equines were reported in Loulé (Algarve) with some animals testing positive by serology [62].

It is likely that the main reason why none of the human or equine reported infections in Portugal have so far been confirmed by RT-PCR is due to diagnosis and screening being performed well past the viraemic phase. This might also explain why obtaining sequencing data related to WNV Portuguese infections has not been successful. To date, a single study has been published on the local phylogenetics of WNV, with sequencing solely obtained from mosquito pools (**Table 4**) [60]. Such sequences, based on a short segment of the NS5 gene, demonstrated a close genetic relationship with past WNV lineages circulating in the Mediterranean basin (France, Morocco and Italy), but fell short in providing definite conclusions on viral diversity, phylodynamics and persistence in the country.

When summarized, the past evidence for the circulation of WNV in Portugal is mostly restricted to reports from the southern regions of the country, in particular the districts of Faro, Beja, Setúbal, Évora, Lisboa, Portalegre and Santarém (**Figure 1, Table 2**). Such reports include evidence from avian species (e.g. *Ciconia ciconia, Strix aluco, Accipiter gentilis*), equines, mosquitoes (e.g. *Culex pipiens, Culex univittatus*) and humans, achieved via different confirmation methods. It is unknown whether the observed WNV circulation in the southern regions, compared to the northern regions, stems from sampling bias, or from disparate ecological conditions dictating WNV epidemic activity. Sampling bias could be due to large differences in regional biodiversity and population sizes of sentinel species (mosquitoes, birds, equines) across the country. Indeed, there is evidence that the equine population is larger in the south [66], while the population sizes and diversity of birds in the north and south are largely unknown for many resident and migratory species. To the ecological point, climate is a major driver of the population dynamics of mosquito and avian species participating in the zoonotic cycle of WNV. According to the Köppen climate classification, Portugal presents a north-south divide in climate types, with the northern region classified as Temperate Mediterranean, and the southern region as Warm Mediterranean. A lack of past studies exploring the effects of climate on WNV epidemiology in Portugal has, however, not allowed for much speculation on its role in the seemingly restricted circulation of the virus to the south of the country. Considering the possible sampling bias and lack of the above mentioned ecological and genetic information, and with other factors still to be researched in the national context, there is currently no consensus on where and whether WNV circulates in Portugal sporadically or endemically.

**Figure 1.**
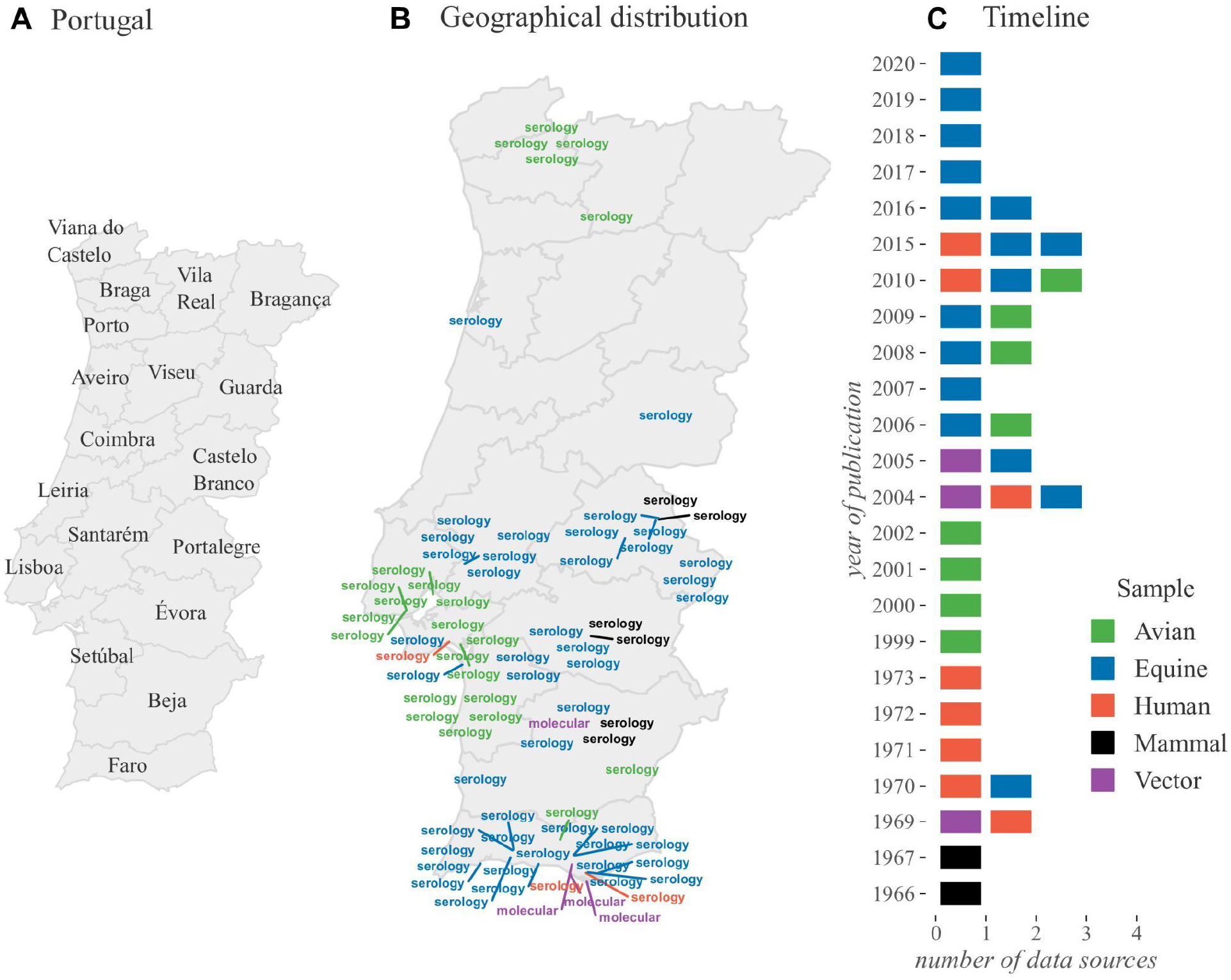
Past evidence of WNV circulation in Portugal. Summary of data sources (articles, national reports) on various types of evidence for the circulation of WNV in Portugal. Panel A presents the official (district) divisions of Portugal. Panel B presents the geographical region of each data source (latitude-longitude). Positioning is only approximate, given that some reports refer to regions of different areas. For data sources referring to large areas (e.g. districts), the capital city of the region was used. Not all sources are mapped, e.g. not reporting where positive samples were found. Mapped words represent the type of evidence reported, aggregated into two categories: molecular, for PCR, isolation; and serology, for HI, VNT, ELISA. Panel C shows a timeline of the number of data sources per year. The color scale (on the right) represents the different types of samples reported, and refers to both panels. Not all years between 1966 and 2020 had data sources. Detailed summary of data sources in **Table 2**.

### 3.2 West Nile virus vectors in Portugal

To date, very few articles in the research literature have provided geo-temporal data on the population size and dynamics of mosquito species relevant for the transmission of WNV in Portugal (**Table 3**). We obtained REVIVE *Culex pipiens* data collected between 2017 and 2019 across Portuguese counties (**Figure 2**). The methods of collection (e.g. type of trap used), as well as resources and regional objectives, varied by year, as described in detail in the published reports [29]. Summarizing the data revealed that both sampling coverage and the total number of yearly collections per county varied in time, both for adult (**Figures 2A-C**) and immature (**Figures 2E-G**) forms of this mosquito species. Moreover, we noticed that even in the presence of many project-related factors affecting sampling, *Culex pipiens* was found to be present across the country (**Figures 2DH**). Since *Culex pipiens* is considered the main vector for the possible transmission of WNV in Portugal [67], these findings stand in contrast with the north-south divide of WNV circulation found in the data review from the literature and public databases (**Figure 1, Table 2**).

**Figure 2.**
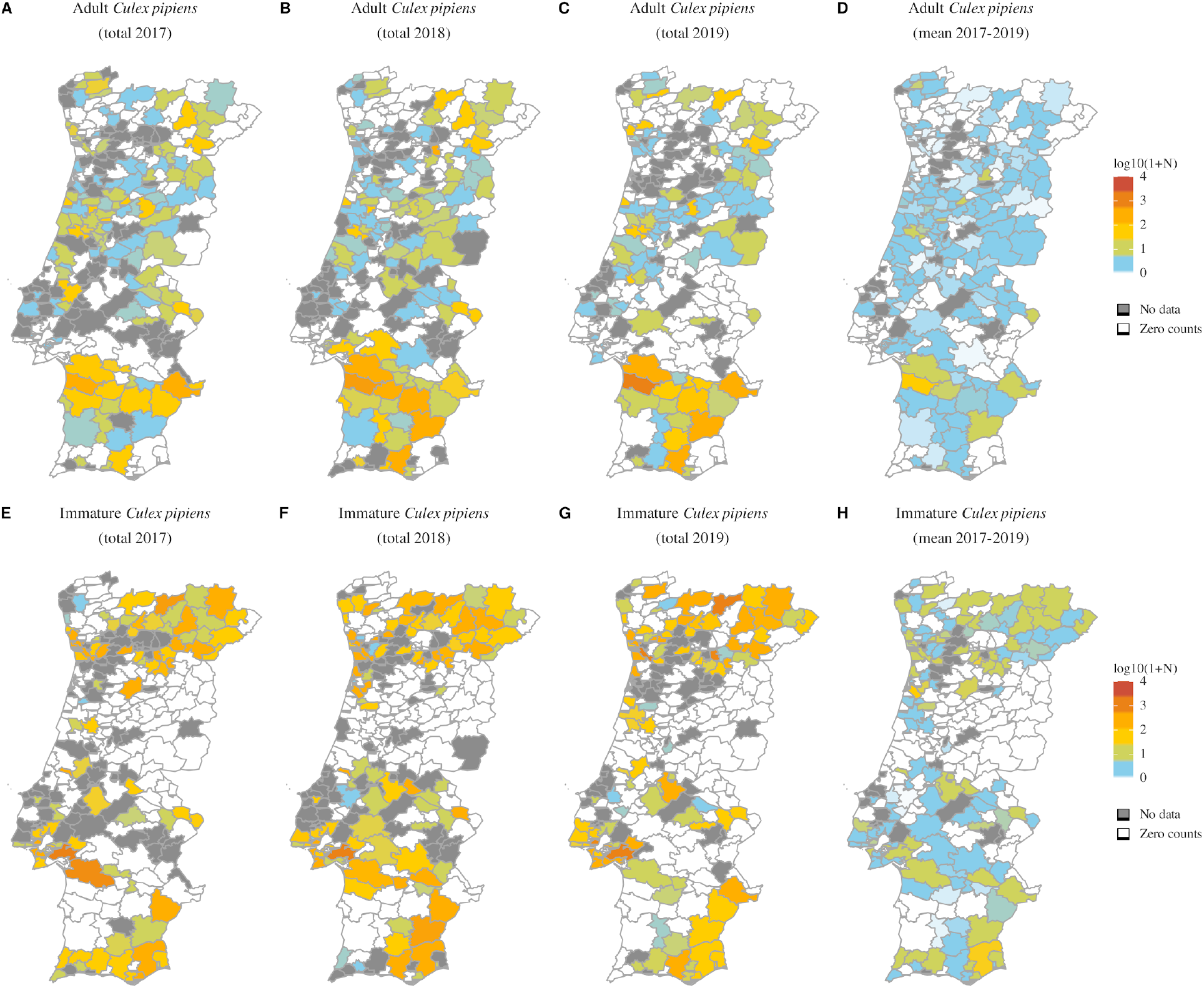
REVIVE Culex pipiens counts in Portugal (2017-2019). Borders represent Portuguese counties, the level at which Culex pipiens data are reported. In panels A-C and E-G, counties are colored according to the yearly observations (transformed to log_10_ (counts + 1)) of adult and immature mosquitoes (respectively). In panels D and H, counties are colored by the yearly mean observations (transformed to log_10_ (mean(counts) + 1)) of adult and immature mosquitoes (respectively). Counties for which there was zero counts are coloured in white. Counties for which collections were not performed are in grey.

### 3.3 West Nile virus transmission potential in Portugal

In this study we use a climate-driven suitability index (termed index *P*) to estimate the, geo-temporal transmission potential of WNV in Portugal. This index estimates the l transmission potential of each adult female mosquito in the zoonotic reservoir, and can be interpreted as a proxy for the risk of spillover to the human and equine populations (see **2.3 Transmission Suitability** for details). Typically, climate-driven suitability indices are validated by quantifying geo-temporal correlations between time series of the index and suspected or confirmed infections. The index used here was previously validated for WNV in Israel [43], and for other viruses elsewhere, with respective modifications [13,36,39]. However, WNV infection time series do not exist for Portugal due to the absence of reported chains of transmission, and hence we resort to index *P* as proxy for WNV transmission (**Figure 1, Table 2**).

We thus first compared the only available time series data related to WNV transmission in Portugal, which included the newly estimated index *P* and regional *Culex pipiens* population sizes from the REVIVE initiative (**Figure 3**). Given the north-south disparity in past evidence for the circulation of WNV (**Figure 1**) and the widespread geographical distribution of *Culex pipiens* (**Figure 2**), we compared directly two of the most northern and southern districts of the country. Although the index *P* measures transmission per mosquito and not population size, we found the index to present peaks and troughs coinciding in time with those of the mosquito population sizes, both in the south (**Figures 3AC**) and north (**Figures 3BD**). Apart from the previously demonstrated potential for the index *P* to be correlated with WNV human infections [43], these results thus suggest that (at least in Portugal) the index can also be used to estimate the seasonal timing of variation in *Culex pipiens* population sizes.

**Figure 3.**
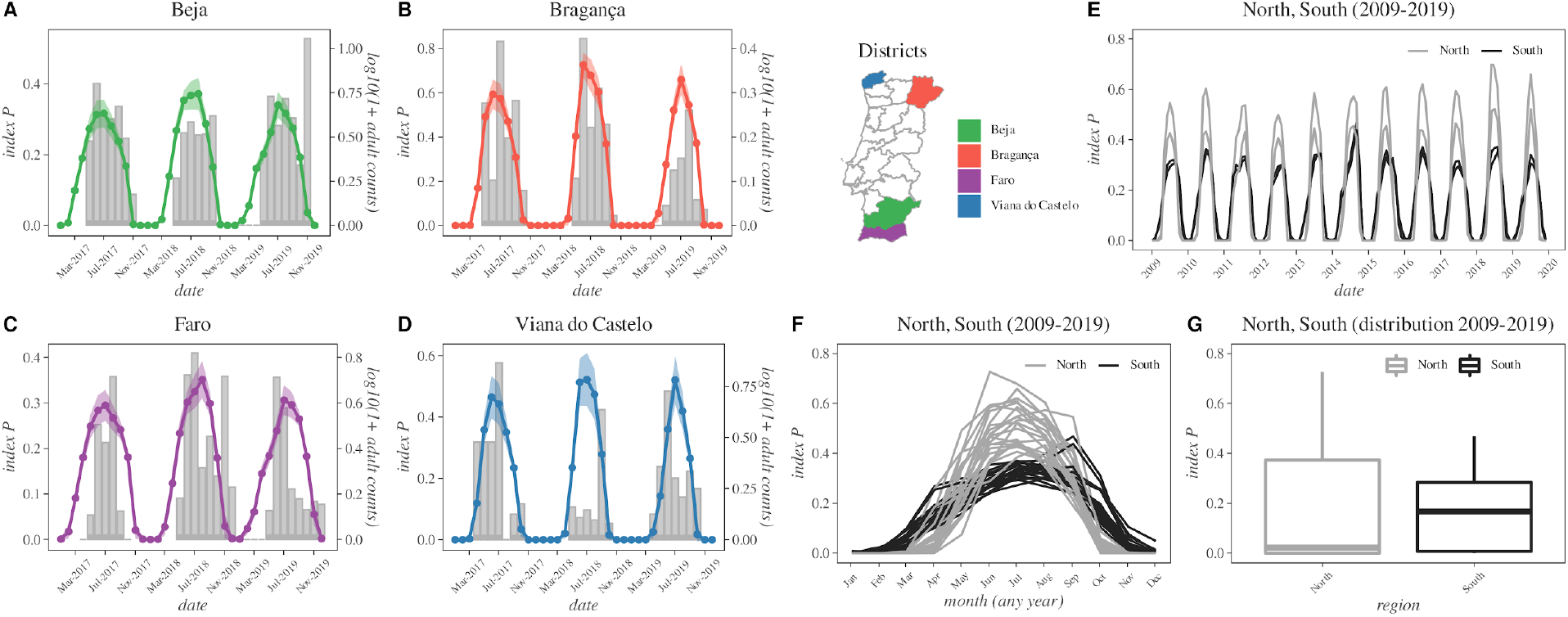
Monthly REVIVE Culex pipiens counts in Portugal versus estimated transmission potential index P (2017-2019). Panels A-D present the monthly time series for log10[1+mosquito population size] (grey bars) and mean (colored lines) and standard deviation (coloured areas) for four districts: two from the south (A for Beja in purple, C for Faro in purple) and two from the north (B for Bragança in red, D for Viana do Castelo in blue). The map panel presents all districts, coloured by their location. Panel E shows the mean index P for all districts 2009-2019 (North in grey and South in black). Panel F shows the seasonal index P per month from panel E independently of year. Panel G is the distribution of index P across the period 2009-2019. For the estimation of the index P, the monthly mean of the climate variables was used per district.

The patterns in **Figures 3A-D** further suggested several characteristics of the typical season of WNV transmission in Portugal, as informed by both mosquito population size and estimated transmission potential. Seasons appeared to occur between the months of March and November of each year, often peaking in the summer and reaching minimum transmission potential values during the winter months. We calculated the peak timing for each region using the index *P* time series of all counties, and found that historically peaks occur in July, although on average 12 days later in the south compared to the north (t-test, *P* < 0. 01, see **2.4 Analysis of peak suitability timing** for details). When comparing the northern and southern districts for a longer time period (2009-2019, **Figures 3EF**), we found that seasonal variation in index *P* typically presented higher values during the summer months in the north compared to the south (≈ 35%higher peaks, t-test *P* < 0. 001. However, compared to the northern districts, the southern districts presented wider seasonal waves, with transmission potential increasing earlier in the spring and remaining non-zero well into late autumn (**Figure 3F**). The higher values of *P* in the north, suggesting higher summer transmission potential compared to the south, contrasted with the existing evidence for WNV circulation (**Figure 1**). However, the long-term time distribution of index *P* seasonal waves in the north was skewed, with lower medians than in the south (**Figure 3G**), potentially supporting the north-south divide found in past evidence for WNV circulation in the country (**Figure 1**).

To explore in detail the regional differences in transmission potential across the country, we next estimated the index *P* per available latitude-longitude coordinate in our climate dataset for the time period between 2016 and 2019 (**Figure 4**). Yearly summary statistics per latitude-longitude coordinate revealed a north-south divide similar to the one presented by past evidence of WNV circulation in the country (**Figure 1**). In particular, we found the southern region of Portugal to universally present higher median yearly index *P* when compared to the north (**Figures 4ACEG**). However, the geographical ‘boundary’ between north and south, in terms of WNV transmission potential, changed over the years (compare e.g. **Figures 4A** to **4C**). This output showed that, in accordance to previously described geo-temporal patterns for other mosquito-borne viruses [36] and WNV [43] in different countries, natural climate variations strongly influence both inter- and intra-yearly variation in transmission potential. When quantifying the number of months within a year for which the estimated transmission potential was not zero, we again found a clear division between the north and south (**Figures 4BDFH**). Regions in the south and south-west typically spent nine or more months of the year with non-zero transmission potential, contrasting to regions in the north and north-east which spent only half or less of the year with non-zero transmission potential. Both the higher yearly median and longer seasonal waves in the south compared to the north, could thus be seen to support the mentioned north-south divide in past evidence for WNV circulation in Portugal (**Figure 1**), hinting that the south may provide particularly favourable conditions for the virus.

**Figure 4.**
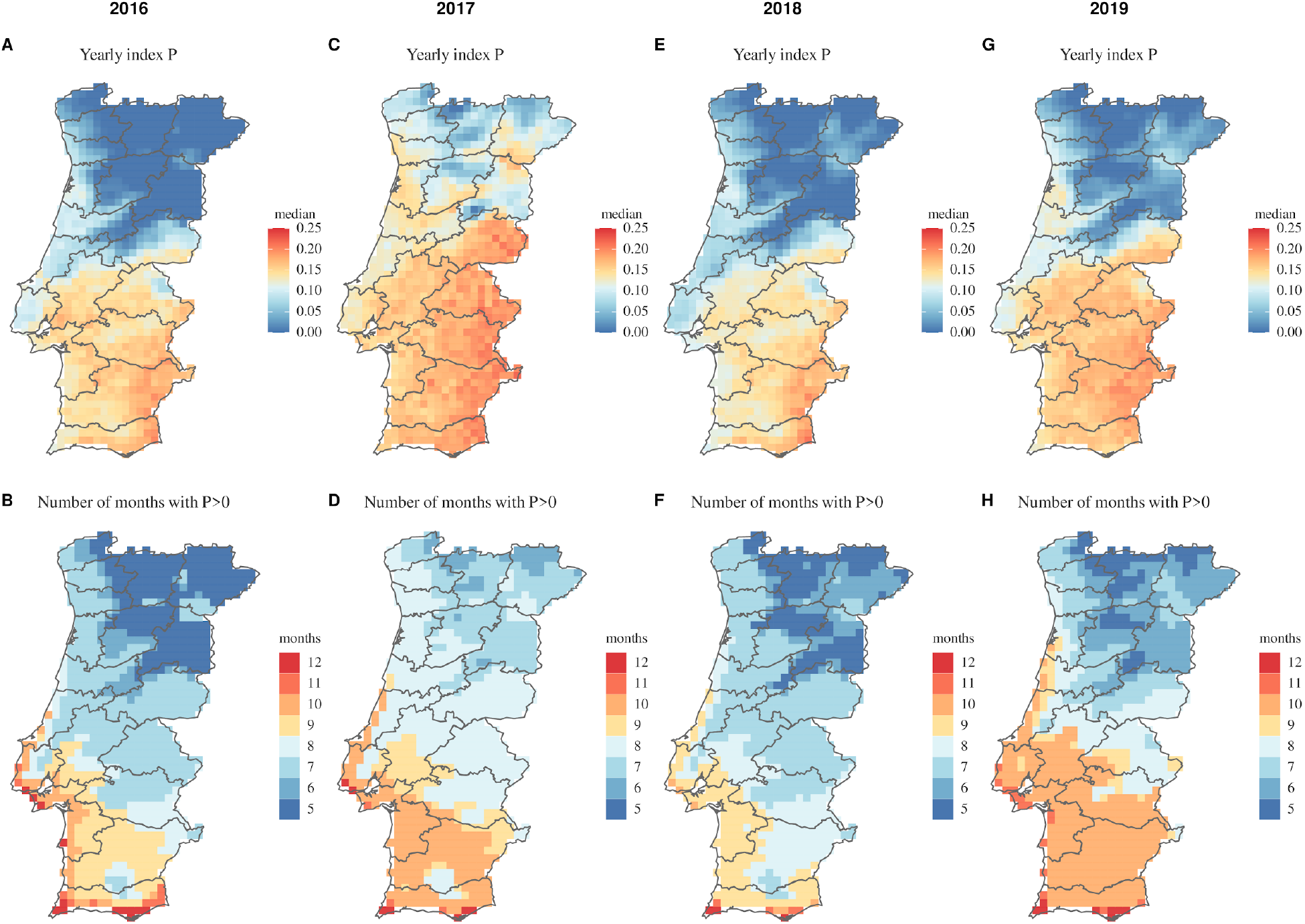
Estimated WNV transmission potential across Portugal (2016-2019). Maps present summary statistics of estimated index P (WNV transmission potential) at the spatial resolution available in the climate dataset. Panels A, C, E and G present the yearly median index P per coordinate according to the presented color scale on the right. Panels B, D, F and H present the number of months during each year for which the estimated index P per coordinate was above zero, according to the presented color scale on the right. Boundaries in dark grey represent districts.

### 3.4 Long-term WNV suitability and climatic trends in Portugal

Estimating transmission potential of a pathogen via the index *P*, while informed by climatic variables, provides means to reconstruct historical trends when long-term climatic data is available. We thus used the past 40 years of annual climate data per Portuguese county to explore long-time trends in climate variables and estimated WNV transmission potential, using linear quantile mixed models (see **2.5 Analysis of long-term climate and suitability trends** for technical details).

We found that all four explored variables presented significant differences between the north and south regions of the country (**Figure 5**). Among these, precipitation had the smallest change over time, with a yearly decrease of ≈−1.2 ⨯ 10^−6^ in the north, and increase of ≈ 4.8 ⨯ 10^−6^ in the south (**Figure 5A**). Following the Köppen climate classifications, temperature was historically warmer in the south compared to the north. There were similar yearly increases of ≈ 0. 041 (north) and ≈ 0. 039 (south) Celsius per year between 1981 and 2019, equating to cumulative changes of ≈ 1. 55 and ≈ 1. 44 degrees for the north and south, respectively (**Figure 5B**). As the country warmed, it also became dryer, with an accumulated decrease in relative humidity of ≈ 2. 8% in the north and ≈ 1. 6% in the south (**Figure 5C**). Such historical climate trends translated into an yearly increase in estimated transmission potential for both regions (**Figure 5D**). The index *P* had an yearly increase of ≈ 0. 00097, accumulating a change of ≈ 0. 038 between 1981 and 2019. During this period, the south had an absolute difference of ≈ 0. 083 in estimated transmission potential compared to the north.

**Figure 5.**
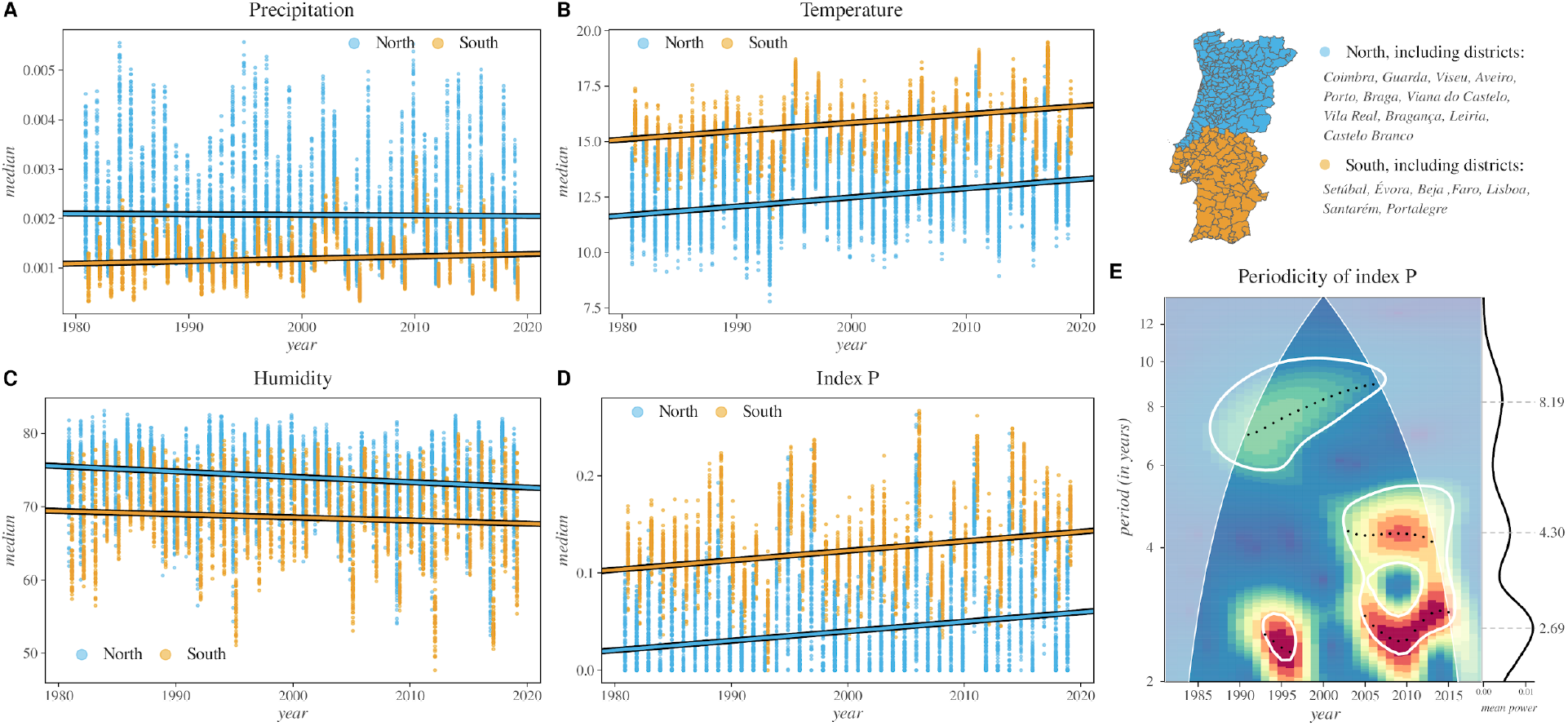
Long-term trends and periodicity of transmission potential of WNV and climate across different regions of Portugal. Panels A, B, and C present the median of climatic variables among counties (points) within the north (blue) and south (orange) regions defined by the set of districts listed in the map subpanel (on the right). Panel D presents the same as A-C but for the index P (transmission potential). Panel E presents the cross-wavelet transform of the median yearly transmission potential between north and south time-series. Significant periodic components are circled in white (p − value < 0. 01), where the heat map denotes power with peaks indicated by black dots (wavelet ridges) and mean power throughout time shown in the right subplot.

As described in **Figure 3**, the northern and southern regions presented an evident yearly periodicity in climate-driven transmission potential, with cycles being stable throughout the years and in phase between the regions. To explore the possibility of periodicity beyond the 12 month cycles, we performed wavelet analysis on the north and south yearly median of transmission potential (see **2.6 Analysis of long-term suitability periodicity** for details). The analysis revealed longer periodic components in both regions, which were seen to change between 1981 and 2019 (**Figure 5E**). Before 2005, periodicity was mostly found in ≈ 8 year cycles, although a strong 2-3 year component was present around 1995. From 2005 onward, periodicity shifted to become strongly dominated not by one but two components at 2-3 and 4-5 years. The presence of such quasi-periods indicates a Moran effect [68], i.e. that climate synchronizes transmission potential across large geographical areas in Portugal beyond the classic intra-year, summer winter cycles. Finally, the significant shifts in the periodic components over the years do not support an historically stable influence of climate, but are rather suggestive that inter-year instability is an expected feature of WNV transmission potential in Portugal.

## 4 Discussion

Recently, several European countries in the Mediterranean basin have quickly progressed from sporadically reporting WNV infections to reporting yearly epidemic activity affecting both humans and equines (e.g. Spain, France, Greece [22]). To date, sufficient evidence has accumulated to support ongoing circulation of WNV in Portugal. However, despite Portugal sharing similar climate types and bird migration routes with nearby countries that have experienced WNV introduction and local dissemination, no events of sustained epidemic activity have been reported. Without such reports, the perception of the public health importance of WNV in Portugal remains minimal. Although several studies have focused on describing WNV human, animal and mosquito infections in the country, its epidemiology remains largely uncharacterised and poorly understood. In this study we have generated an historical, comprehensive perspective of the virus in Portugal using a combination of existing data and climate-informed theoretical approaches.

We collated existing data from the research literature, national databases and recent serological samples in equines representative of the time period between 1966 and 2020. Recent data revealed that *Culex pipiens*, considered to be the most suitable species for WNV transmission in Portugal, is present across the entire country. In contrast, we found that the vast majority of WNV-related evidence has been reported in the districts of Faro, Beja, Setúbal, Évora, Lisboa, Portalegre, and Santarém. This group of districts creates a clear north-south divide at the ≈ 39. 5 latitude axis of the country, suggesting that past or ongoing WNV circulation is restricted to the south of Portugal. Coincidently, such a divide is representative of the known geographical separation of the two Köppen climate classifications that exist in the country, with the north being classified as Temperate Mediterranean and the south as Warm Mediterranean.

Climate has long been accepted as a major driver of WNV epidemic activity and dissemination, given its effects on both the mosquito and avian species’ population dynamics. To consolidate and complement existing data, we explored the effects of local climate variation on theoretical estimations of WNV transmission potential over the years. We estimated that, similarly to other countries in the northern hemisphere, WNV transmission seasons across the country tended to occur between March and November (Spring, Autumn), often peaking in July (Summer). The northern districts presented higher WNV transmission potential during the summer months, compatible with the expectation that WNV potential peaks at intermediate temperatures of 23-26 celsius [19]. Indeed, during the summer months (June-August, 1981-2019), the average temperature of the northern districts was above 26 celsius only ∼3.1% of the time ; as opposed to ∼43% in the southern districts Hence, the generally warmer climate in the south may be detrimental for WNV transmission in the peak of the summer. Additionally, we found that the inter- and intra-year median transmission potentials were substantially higher in the south when compared to the north. This implies that the south presents longer seasons suitable for WNV transmission, including the spring and autumn months; while the suitable seasons in the north are mostly restricted to the summer months. The importance of longer seasons suitable for WNV transmission rests on prior data: *Culex* mosquitoes from North America and Europe have been observed to shift their feeding preferences by late summer and early autumn, coinciding with a decrease in abundance of particular avian species and increases in reported infections [69,70]. Together, these differences between the north and the south of Portugal support the existing north-south divide in the evidence of WNV circulation.

In light of the ongoing discussion of the effects of climate change on the long-term epidemiology of mosquito-borne viruses (including WNV), we explored historical climate variations and the corresponding WNV transmission potential in Portugal. Differences between the north and south were clear and remained similar over the past 40 years. In general, we presented evidence that Portugal has slowly changed towards a warmer and dryer climate over this period. Temperature presented an especially large change across the years, with a median increase of +1 degree Celsius. Temperature remains a critical factor in mosquito-borne epidemiology, often positively affecting a large number of viral and entomological traits that favour transmission potential. Key examples are the positive relationship of higher temperatures with a shorter viral extrinsic incubation period, longer adult mosquito life-span and shorter aquatic mosquito development [19,33,71]. Following the aforementioned historical trends in climate, we also found a small but continuous increase in transmission potential over the years. Our analyses also suggest that recent changes in climate, since 2005, appear to have introduced unstable inter-year variations in transmission potential, the practical consequences of which are difficult to assess.

The data and modelling outputs presented include some limitations. For example, the geotemporal patterns of transmission potential reflect solely the contribution of natural climate variation. Our estimations do not take into consideration other factors that could affect WNV transmission suitability in Portugal. These may include the geographical distribution of adequate terrain and vegetation (e.g. wetland, marshland) that favour avian-vector mixing and their proximity to human populations, or geo-temporal hotspots for inbound migratory avian species, vector and avian population sizes, biotype composition of the *Culex pipiens* populations, etc. Such factors should be the basis of future research extending the methods and analyses presented in this study, when such data becomes available at adequate resolutions. The original resolution of the climate data has also restricted some of our outputs. Namely, the resolution of our maps may have missed small-area regions of importance, and we were only able to estimate the duration of seasons (non-zero transmission potential) in the scale of months. The collated empirical data of past WNV circulation may be another source of limitations. For instance, some evidence was impossible to map geographically, given deficiencies in reporting (e.g. no region, or non-specific regions e.g. “south” or “litoral”). It also remains unknown if the data includes a sampling bias towards the south of the country, which could partially explain the seemingly north-south divide in WNV circulation. This bias could arise from the reactive nature of surveillance initiatives responding to reports of symptomatic disease in sentinel species which may be distributed differently across the country, or alternatively, due to differences in WNV awareness among farmers and veterinarians.

The large European WNV epidemic of 2018 provided key opportunities to assess existing WNV surveillance and public health mechanisms. Countries implementing cross-sectoral and cross-disciplinary early warning systems based on One Health surveillance (vector, avian, equine and human) consistently reported positive impacts of such infrastructures [21]. Common areas for possible improvement included investment and sustainability of mosquito surveillance, and improved information and media management. In Portugal, mosquito surveillance remains an exception in an otherwise fragmented and insufficient WNV surveillance infrastructure. Key improvements would include sustainability of laboratory capacities and their timely responses, active One Health surveillance beyond the mosquito, and investment in awareness and training across public health domains and the general public, in particular in the south of the country.

## 5 Conclusions

Our study provides a comprehensive data-driven WNV perspective, and a first spatio-temporal assessment of the contribution of local climate to the theoretical transmission potential of the virus. The ubiquitous countrywide oscillations in transmission potential between summer and winter months demonstrate that climate can couple WNV seasons throughout Portugal, independently of the absolute transmission potential of each region. Consequently, concerted surveillance and control actions spanning the entire territory will be required in the future to better understand and tackle WNV spread. Past data suggests that WNV circulates mostly in the south of Portugal, infecting birds, equines, mosquitoes, and occasionally humans. In contrast, mosquito data shows that the spatial distribution of WNV-capable species includes all districts, from north to south. Natural differences in climate types between the north and the south contribute to relevant quantitative differences in WNV theoretical transmission potential, supporting the geographical divide observed in molecular and serological data. It is still uncertain if WNV circulates endemically in any region of the country, but sufficient evidence exists to support ongoing seasonal circulation. The south of Portugal may be more suitable for WNV transmission, but the possibility of the north supporting transmission in shorter periods of each year can not be rejected. Portugal is slowly accruing suitability for WNV transmission, and hence a shift from passive to active One Health surveillance will be necessary to prevent and manage future epidemics that may result in human public health emergencies, as reported recently for other countries in Europe. Our results carry implications for future research, surveillance and public health policy.

## Data Availability

All data is deposited in online repositories and links are included in this manuscript.

## 7 Acknowledgements

DSCD was funded by the grant 19/23343-7 and 20/06160-3 within the scope of 15/22308-2 from the São Paulo Research Foundation (FAPESP). JL was supported by a research lectureship by the Department of Zoology, University of Oxford. MG was supported by Fundação de Amparo à Pesquisa do Estado do Rio de Janeiro (FAPERJ). We are grateful to The Ministry of Health / National Institute of Health (INSA) under the National Vector Surveillance Network - REVIVE for supporting this research. We are also grateful to the REVIVE team for the mosquito collection nationwide, and to Dr Patrícia Rosa Ramos Rodrigues for the collection of the reported CSF equine sample. Funding sources had no involvement in the design and interpretation of the presented research.

